# The Effects of Daily Six Major Pollutants on the Risk of Respiratory Disease-Related Emergency Ambulance Calls: A Six-Year Time Series Study

**DOI:** 10.1101/2025.03.24.25324565

**Authors:** Haotian Jiang, Zhifeng Zhang, Li Peng, Wenjie Lu, Jiayue Zhu, Yan Hu, Xu Liu

**Author notes:** Correspondence (X.L.). These authors contributed equally to this work (Haotian Jiang, Zhifeng Zhang, Li Peng). (H.J.); (W.L.); (Y.H.). (J.Z.). (Z.Z.). (L.P.).

## Abstract

**Background:** In recent years, the impact of air pollution on the emergency departments of medical institutions has become increasingly evident. Emergency Ambulance Calls (EACs), compared to traditional health indicators such as mortality and hospitalization rates, provide a more direct reflection of the short-term effects of air pollution on public health. This study aims to explore the short-term association between the daily average concentrations of six major pollutants (PM_2.5_, PM_10_, SO_2_, NO_2_, CO, O_3_) and EACs related to respiratory diseases in the central urban areas of Shanghai. Methods: The Generalized Additive Model (GAM) was used to estimate the excess relative risk (ERR) of each pollutant on EACs at different lag times (0-7 days). Stratified analyses were also conducted based on age, time of day, and season. Results: 122,037 respiratory diseases related EACs were recorded during the study period. In different lag-day models, each interquartile range increase in pollutant concentration was associated with the highest single-day lag excess risk of EACs on the 6th day for all six pollutants, except for O3, which peaked on the 3rd day. The study found that individuals aged 65 and above are a vulnerable population to exposure. Specifically, in spring, PM_2.5_ on the 6th day of single-day lag was associated with a 3.19% (95% CI, 1.48-4.93%) increase in all-day EACs risk; PM_10_ on the 7th day of cumulative lag was associated with a 4.98% (95% CI, 1.35–8.74%) increase in daytime EACs risk; and O_3_ on the 3rd day of single-day lag was associated with a 3.60% increase in daytime EACs risk among the elderly (95% CI, 1.19–6.06%). Conclusion: This study indicates that even under the national ambient air pollutant concentration limits, air pollution could still serve as significant triggers for acute respiratory disease exacerbations. It is recommended that stricter air pollution control and early warning policies be implemented to reduce the occurrence of respiratory disease-related emergencies.

## 1. Introduction

It is well known that exposure to air pollution leads to excess respiratory-related morbidity and mortality [1,2]. In the context of limited pre-hospital emergency resources, this poses a significant public health burden on the healthcare system. Over the past decade, an increasing number of epidemiological studies have examined the relationship between short-term exposure to air pollutants and the incidence of respiratory diseases [3,4]. Additionally, some studies have begun to focus on the short-term health impacts of air pollution exposure on the public, specifically the variation in Emergency Ambulance Calls (EACs) [5, 6]. The daily average concentrations of the six monitored pollutants are important indicators for assessing the health impacts of air pollution. These concentrations may have different effects on EACs across seasons, as well as during the day or night [7]. Investigating the various impacts of different pollutants can help reduce the relative risk of exposure. Although previous studies have examined the relationship between daily average concentrations of air pollutants and emergency calls, their limitations cannot be overlooked, such as small sample sizes, short study durations, exclusion of all six monitored pollutants, and a lack of focus on respiratory-related health impacts [6, 8].

In response to the growing air pollution crisis, the World Health Organization (WHO) has formulated a series of policies to effectively mitigate air pollution and has established a set of air pollution limits in the *WHO global air quality guidelines* [9]. But still a study found that even when air quality meets the new WHO guideline threshold (15μg/m^3^), an increase in PM2.5 concentration remains associated with a heightened risk of EACs for respiratory diseases [10]. Asia is one of the regions with the most severe air pollution worldwide. As the largest country of Asia, the Chinese government has implemented a series of policies to vigorously control air pollution, which have contributed to a reduction in pollution levels to some extent. Additionally, corresponding limit values have been set for the six major pollutants due to *Ambient air quality standards* implanted on 2016-01-01 by the Ministry of Environmental Protection of China [11]. As one of the largest cities in Asia, Shanghai still needs to strengthen research on pre-hospital emergency demand related to air pollutants, especially in the context of ongoing urbanization and rapid climate change.

Considering the limited evidence linking six major pollutants with well-defined EACs, along with the absence of classification for potential patterns (such as seasonal variations and daytime/nighttime effects) that could influence respiratory-related EACs and positive flaw of six major pollutants limit values, it is crucial to focus on addressing these research gaps in this study. Thus, we conducted a generalized additive model (GAM) to examine the predominant lag for each air pollutant and determine new pollutant limits based on the exposure-response curve; then, we adopted a stratified analysis to explain the specific mechanisms by which each pollutant affects respiratory disease-related EACs. Furthermore, we hope to explore new limits through this study to better support policy making related to air pollution control.

## 2. Materials and Methods

### 2.1 Study Participants

The Shanghai Emergency Medical Center (SEMC) established a system in 2013 composed of the 120 Emergency Medical Services (EMS), which includes 119 emergency medical institutions and 166 emergency stations. We utilized vehicle dispatch records from 54 emergency sub-stations in the central urban areas of Shanghai, provided by the SEMC, covering the period from January 1, 2016, to December 31, 2021. The study area encompassed seven administrative districts in the central urban area, as well as two streets in the Lujiazui area of the Pudong New District. The inclusion criteria for the study data were as follows: (1) No missing values for dispatch times; (2) Calls originating from the central urban districts of Shanghai, including Huangpu, Xuhui, Changning, Jing’an, Putuo, Hongkou, Yangpu, and the Pudong New District; (3) Dispatch times within the period from January 1, 2016, 00:00:00 to January 1, 2022, 00:00:00; (4) Disease categories including “respiratory system.” Given that respiratory diseases are generally common conditions, the misdiagnosis rate in the data is relatively low. A total of 23 items, including emergency call address, administrative district, dispatch location, dispatch time, basic vital signs of the patient, medical history, preliminary impression, and others, were recorded.

### 2.2 Ethics

This study has received approval from the ethics committee of the Naval Medical University. All the participants voluntarily agreed to take part in our survey. During this study, the data provided by the Shanghai Emergency Medical Center had been thoroughly anonymized, with all directly identifiable personal information (such as names, ID numbers, and exact residential addresses) removed. Throughout the data collection and analysis phases, authors had no access to any information that could directly identify individual participants, nor could individual identities be inferred from the available data.

### 2.3 Outcome Ascertainment

Our primary outcome was respiratory EACs. In this study, the respiratory disease-related diagnoses primarily encompass chronic bronchitis, chronic obstructive pulmonary disease (COPD), and obstructive emphysema. As patients transported to the hospital require further diagnostic evaluation to establish a definitive diagnosis, the vehicle dispatch records do not include specific ICD disease classification codes.

### 2.4 Air Pollutant Exposure Ascertainment and Covariates

In this study, we obtained daily air pollution data and daily meteorological data for the central urban areas of Shanghai from the Shanghai Environmental Monitoring Center. Based on the results of Spearman correlation analysis and relevant literature (Figure S1), when constructing the Generalized Additive Model (GAM), we incorporated the two meteorological variables most strongly associated with each pollutant to more accurately estimate the impact of meteorological variables on different pollutants. Issues of multicollinearity have been eliminated (i.e., ensuring that highly correlated meteorological variables were not included as covariates simultaneously). Finally, we included combinations of wind speed and minimum temperature, relative humidity and minimum temperature, mean temperature and sunshine duration, relative humidity and minimum temperature, relative humidity and minimum temperature, and wind speed and minimum temperature as covariates for each pollutant (PM_2.5_, PM_10_, O_3_, SO_2_, NO_2_, CO). Seasonal divisions in the study were determined based on the relevant indicators in the *Division of climatic seasons* implanted on 2023-02-01 by the Standardization Administration of China (SAC) [12]. Specifically, in the 5-day moving average temperature series, the initial determination of the climate season start date was made based on the first consecutive 5 values of the moving average sequence that met the threshold of a specific climate season indicator in the corresponding 9-day daily average temperature series, according to the following criteria:

- Spring: The first date with a daily average temperature greater than or equal to 10°C.
- Summer: The first date with a daily average temperature greater than or equal to 22°C.
- Autumn: The first date with a daily average temperature less than 22°C.
- Winter: The first date with a daily average temperature less than 10°C.

The day prior to the start date of a given climate season is considered the end date of the previous climate season for that year.

### 2.5 Statistical Analysis

We used the Generalized Additive Model (GAM) to estimate the short-term association between each pollutant and daily respiratory-related calls. According to previous related studies, The model employed a quasi-Poisson distribution to account for the overdispersion in daily emergency call counts [13]. In the model, we controlled season variables and used smooth spline functions to control for long-term trends. We selected the model specification and degrees of freedom (df) for the smoothing functions based on the Akaike Information Criterion (AIC) values of the model. Specifically, when wind speed and minimum temperature were used as covariates, df were set to 3 and 5, respectively. When relative humidity and minimum temperature were used as covariates, the df were set to 5 and 3, respectively. When mean temperature and sunshine duration were used as covariates, the df were set to 3 and 6, respectively. The main model is as follows:

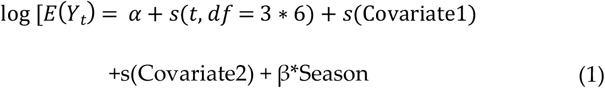

In the model, *E*(*Y*_*t*_)represents the respiratory disease-related emergency calls on day *t, α* is the intercept term, and in the actual model, it corresponds to the concentration of each pollutant at different lag days. The term *s*() denotes the smooth spline function, and in the actual model, cubic splines were chosen for the connection. The *t* variable describes long-term effects, while *β* variables are the regression coefficients, which were implemented in the actual model by treating the season variable as a factor variable.

Based on the previous study, for example, a study that focusing on the health effects of air pollution also conducted in Shanghai [14]. We estimated the lag effects from the same day (lag0) to 7 days prior (lag7). We also examined the cumulative effects generated by the moving averages of the same day and the previous 7 days (lag01-06, respectively). The excess relative risk (ERR) of daily EACs was estimated for each interquartile range increase in the concentration of each pollutant. The calculation method for ERR is as follows:

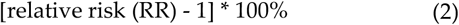

RR values are obtained from the GAM model.

### 2.6 Stratified Analysis

Based on the results of the sensitivity analysis, we selected PM_2.5_ at a single-day lag of 6 days, PM_10_ at a single-day lag of 6 days and a cumulative lag of 7 days, O_3_ at a single-day lag of 3 days, and SO_2_ at a cumulative lag of 7 days for inclusion in the stratified analysis. Subgroups were defined based on the preliminary descriptive analysis, specifically categorizing age into under 65 years and equal or more than 65 years, time periods into daytime and nighttime (day: 7:00-20:00; night: 20:00-7:00), and seasons into spring, summer, fall and winter. Exposure-response curves were generated for each subgroup in the stratified analysis to reveal the risk of acute respiratory disease exacerbations among vulnerable populations during seasons with higher exposure risks.

## 3. Results

Between 2016 and 2021, a total of 1,095,195 EACs occurred in Shanghai. Among these, 122,037 calls were related to respiratory diseases. With the exception of a decline in 2020 due to the global COVID-19 pandemic, the number of these calls exhibited an annual increasing trend, with 16,480, 17,298, 19,078, 21,794, 21,510, and 25,877 calls in each respective year. Before 2020, the proportion of elderly individuals among EACs was dominant, accounting for approximately 84% of calls in the overall population. However, following the outbreak of the COVID-19 pandemic at the end of 2019, the proportion of elderly-related calls significantly decreased, while the number of calls from individuals under the age of 65 notably increased. Even in 2021, when the total number of respiratory-related EACs significantly rose, the proportion of calls from individuals under 65 remained stable. The ratio of night-time to daytime calls showed no significant variation, with daytime calls accounting for approximately 70% of the total across all time periods. The proportion of calls from males was slightly higher than that from females, with a consistent annual ratio of approximately 56% and 44%, respectively. Additionally, the average concentrations of air pollutants and meteorological variables are presented in Table 1. As shown in Figure S1, there is a moderate to low correlation between air pollutants and meteorological variables (Spearman coefficient r < 0.80).

**Table 1.**
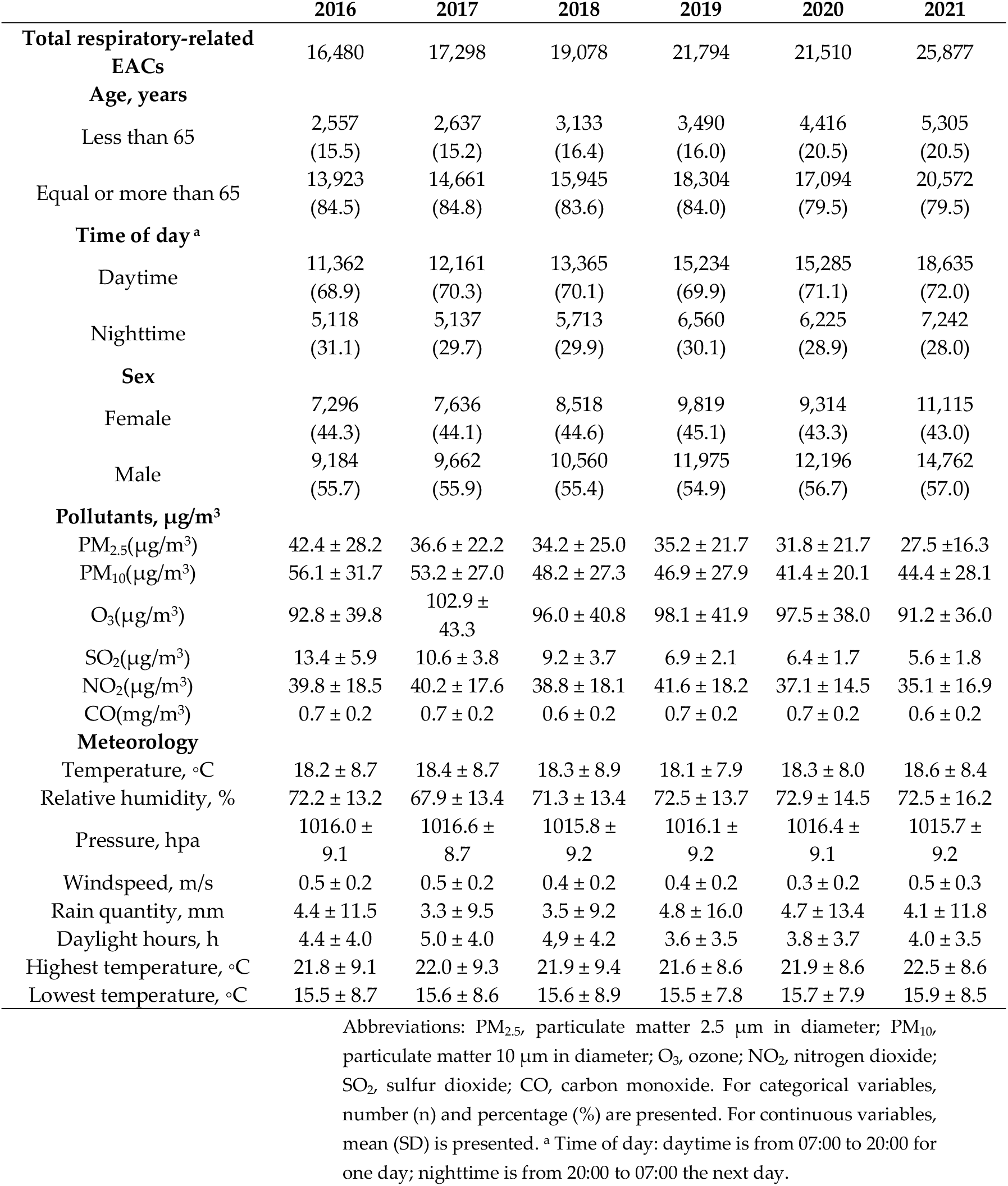
Fundamental features in Shanghai from 2016 to 2021.

The risk of EACs for PM_2.5_, PM_10_, O_3_, SO_2_ and CO exceeded zero before reaching the pollutant limit values. Among these, the concentrations of PM10, SO2, and CO did not even reach the alert threshold (p<0.5, Figure 1). We observed that the relative risk of exposure to PM_10_ and O_3_ increased linearly, while the risk for PM_2.5_, SO_2_, and CO began to decrease after reaching a certain concentration. The relative risk of exposure to NO_2_ remained close to zero until the concentration approached the alert threshold, after which it increased above zero.

**Figure 1.**
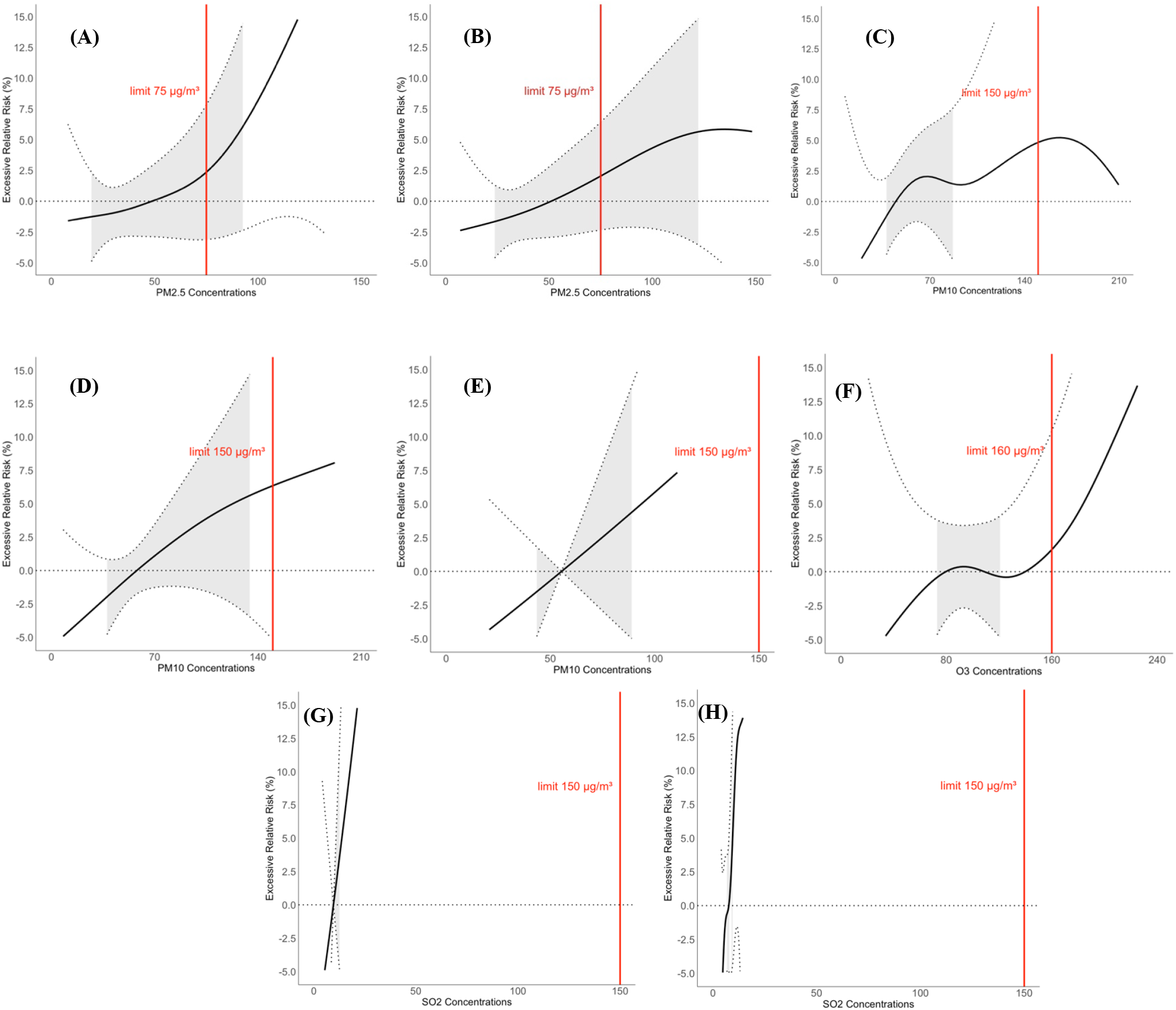
Exposure-response relationship between statically significant findings in stratified analysis and respiratory related disease EACs: **(A)** and **(B)** represents PM_2.5_ on the 6th day of single-day lag during spring and winter, respectively, for the entire day; **(C)** and **(D)** represents PM_10_ on the 6th day of single-day lag during spring and winter, respectively, for the daytime; **(E)** represents PM_10_ on the 7th day of cumulative-day lag during spring, for the daytime; **(F)** represents O_3_ on the 3th day of single-day lag during spring, for the daytime; **(G)** and **(H)** represents SO_2_ on the 7th day of cumulative-day lag during spring and summer, respectively, for the daytime. Abbreviation: All curves represent the EACs for the same age group, specifically individuals aged 65 and above. The black solid lines are the excessive relative risk of respiratory EACs, the red solid lines are the representative of the national regulatory limits, and the dotted lines are the 95% confidence intervals.

In models with different lag periods, we found that each IQR increment in the concentrations of PM_2.5_ (26 µg/m^3^), PM_10_ (31 µg/m^3^), O_3_ (49.25 µg/m^3^), SO_2_ (4 µg/m^3^), NO_2_ (22 µg/m^3^), and CO (0.3 mg/m^3^) was associated with the highest ERR of EACs(1.68%, 95% CI, 0.79–2.58%; 1.30%, 95% CI, 0.42–2.20%; 1.40%, 95% CI, 0.32–2.48%; 1.08%, 95% CI, 0.05–2.12%; 1.31%, 95% CI, 0.23–2.40%; 1.79%, 95% CI, 0.66–2.94%) on the 6th day, except for O3, which showed a peak on the 3rd day (Figure S2, p<0.5). All pollutants exhibited the highest ERR of EACs (1.92%, 95% CI, 0.24–3.63%; 2.50%, 95% CI, 0.82–4.22%; 2.24%, 95% CI, 0.42–4.08%; 2.12%, 95% CI, 0.20–4.07%; 0.19%, 95% CI, −1.58–2.00%; 2.72%, 95% CI, 0.58–4.91%) at the cumulative lag of 7 days (Figure S2, p<0.5).

Based on the highest ERR of each pollutant from models with different lag days, we conducted sensitivity analysis (Table S1). According to the results of sensitivity analysis, pollutant models deemed robust were incorporated into the subsequent stratified analysis (Table 2), which revealed that all included pollutants posed a greater health risk to individuals aged 65 and above. Specifically, PM_2.5_ was associated with an increased risk of EACs throughout the entire day, whereas other pollutants had a more pronounced effect during the daytime. The seasonal effects of different pollutants varied across different lag days. For example, PM_10_ exhibited a higher risk on the 6th day of single-day lag in winter, while its highest risk was observed on the 7th day of cumulative lag in spring. Winter and spring were identified as the two seasons with the highest risk values in the stratified analysis.

**Table 2.** Associations of each IQR increment in PM_2.5_, PM_10_, O_3_ and SO_2_ of different lag days with excessive relative risk of respiratory-related EACs stratified according to age, time of day and season, bold indicates p<0.5.

| Subgroups | PM<sub>2.5</sub>lag6 | PM<sub>10</sub>lag6 | PM<sub>10</sub>lag06 | O<sub>3</sub>lag3 | SO<sub>2</sub>lag06 |
| --- | --- | --- | --- | --- | --- |
| **Age groups, years** |  |  |  |  |  |
| Less than 65 | 1.81 (-0.27, 3.93) | 0.17 (-1.88, 2.28) | -0.30 (-4.09, 3.64) | 1.00 (-1.44, 3.50) | -1.02 (-5.44, 3.60) |
| Equal or more than 65 | **1.63 (0.71, 2.56)** | **1.53 (0.61, 2.47)** | **3.08 (1.31, 4.88)** | **1.49 (0.36, 2.63)** | **2.73 (0.73, 4.77)** |
| **Time of day** |  |  |  |  |  |
| Daytime | **1.68 (0.66, 2.72)** | **1.56 (0.53, 2.59)** | **3.00 (1.05, 5.00)** | **1.45 (0.22, 2.71)** | **2.57 (0.34, 4.84)** |
| Nighttime | **1.55 (0.16, 2.97)** | 0.71 (-0.67, 2.11) | 1.31 (-1.29, 3.98) | 1.27 (-0.43, 2.99) | 1.08 (-1.86, 4.11) |
| **Season** |  |  |  |  |  |
| Spring | **3.19 (1.48, 4.93)** | **1.68 (0.11, 3.27)** | **4.98 (1.35, 8.74)** | **3.60 (1.19, 6.06)** | **7.38 (2.49, 12.51)** |
| Summer | -0.20 (-2.07, 1.72) | 1.21 (-0.66, 3.11) | 2.58 (-0.93, 6.22) | 1.04 (-0.09, 2.19) | **6.08 (1.72, 10.63)** |
| Fall | 0.15 (-2.12, 2.47) | -0.15 (-2.07, 1.81) | 0.97 (-2.65, 4.72) | 0.55 (-3.43, 4.70) | -1.44 (-7.31, 4.80) |
| Winter | **1.37 (0.12, 2.64)** | **1.91 (0.47, 3.38)** | 0.33 (-2.97, 3.75) | 4.37 (-0.31, 9.28) | -0.55 (-3.39, 2.37) |

Subsequently, exposure-response curves (Figure 1) were generated for statistically significant subgroups identified in the stratified analysis. After adjusting for time periods and seasons, it was observed that individuals aged 65 and above exhibited higher excess relative risk upon exposure. However, the seasonal variations of different pollutants varied.

Specifically, for O_3_, the relative risk began to exceed zero as its concentration approached the alert threshold. PM_2.5_ showed a rapid increase in relative risk on the 6th day of single-day lag in spring, with no declining trend even after reaching the alert threshold. In contrast, in winter, the relative risk of PM_2.5_ began to decrease shortly after reaching the threshold. PM_10_ exhibited no significant change on the 6th day of single-day lag in spring, with relative risk remaining close to zero, whereas in winter, its relative risk continued to rise after surpassing the alert threshold, following a trend similar to that observed at a cumulative lag of 7 days.

As for SO_2_, the relative risk at a cumulative lag of 7 days showed a rapid increasing trend in both spring and summer; however, the wide confidence intervals suggest that further discussion is needed to determine its reliability.

## 4. Discussion

Our long-term, large-scale study conducted in Shanghai, China, found a significant association between exposure to PM_2.5_, PM_10_, O_3_, and SO2 during specific seasons and time periods with an increased risk of EACs. Moreover, the risk reached hazardous levels even before these pollutants met the national alert thresholds. This association was particularly evident among the elderly population (*≥*65 years old). These findings are of great significance in the context of global climate change, as they highlight the need to adjust air pollution alert thresholds to mitigate the increase in pollutant-related respiratory disease-related EACs.

Our study utilized daily air pollution data to examine the comprehensive impact of six major air pollutants on respiratory disease-related EACs from the perspective of different lag days. This provides valuable insights for emergency medical departments to proactively prepare for severe air pollution events, optimizing ambulance dispatch and reducing response time delays. By identifying the peak risk values of different air pollutants across various lag periods, healthcare providers can enhance preparedness and response strategies, ultimately improving patient outcomes. Our findings are consistent with a large body of previous research, all of which indicate a positive correlation between daily air pollutant concentrations and the acute exacerbation of respiratory diseases [1, 15-17]. For example, from 2009 to 2017, emergency admission data from a major hospital in Singapore indicated a positive correlation between short-term exposure (within 7 days) to higher concentrations of atmospheric pollutants, including SO2, PM_2.5_, PM_10_, and NO2, and an increased risk of respiratory-related emergency admissions[15]. A three-year study conducted in Delhi, India, demonstrated that short-term exposure (0–7 days) to air pollutants such as PM_10_ and NO2 was closely associated with the worsening of asthma symptoms and an increased rate of acute respiratory emergency visits [1]. In South Korea, a study found that both the cumulative effect and single-day concentrations of PM10 and PM2.5 were positively associated with respiratory disease-related emergency visits. Among the elderly population, the number of emergency visits peaked at a lag of 14 days [16]. The study by Liu et al. in Lanzhou, China, demonstrated a significant association between the concentrations of PM_2.5_, PM_10_, SO2, NO2, and CO and the daily number of emergency visits for upper respiratory tract infections [17].

In analyzing this association, existing studies have primarily focused on the combined effects of multiple pollutants [18], the short-term lag effects of specific pollutants [8] (on an hourly scale), long-term lag effects [14] (0–7 days), and changes in emergency care demand for specific diseases. However, considering the unique geographical characteristics of Shanghai, the limited research specifically focusing on respiratory disease-related emergency care demand, and the greater efficiency of controlling individual pollutants compared to multiple pollutants simultaneously, it is crucial to conduct a comprehensive study on the impact of six major pollutants in Shanghai across different lag periods and seasons on the risk of respiratory disease-related EACs.

In this study, we employed the Generalized Additive Model (GAM) to assess the excess risk of respiratory disease-related EACs associated with the daily average concentrations of six major pollutants across different lag days. This approach allowed us to identify the lag period at which each pollutant posed the highest risk, which was then included in the sensitivity analysis to evaluate model robustness. Previous studies have also utilized similar statistical methods, but they primarily focused on investigating the associations between air pollution and the incidence of respiratory and cardiovascular diseases [19, 20]. For example, Huang et al. studied the impact of air pollution on the spread of COVID-19 in different cities across South America, using a Generalized Additive Model (GAM) to assess the effects of PM2.5 and NO2 on the progression of the pandemic [19]. Tang et al. utilized a Generalized Additive Model (GAM) to quantify the relationship between short-term exposure to air pollutants and the incidence of emergency stroke cases [20].

Our study revealed that in the single-day lag analysis, O3 reached its peak excess risk on the third day, which differs from the patterns observed for other pollutants. This finding is consistent with previous research conclusions [21,22]. Additionally, all pollutants exhibited an approximately linear increasing trend in excess risk across cumulative lag days, which is proved by previous studies [23]. Furthermore, in the exposure-response curve for the cumulative 7-day lag, we observed that NO2 only began to exhibit hazardous effects after exceeding the alert threshold. This indicates that the current NO2 alert threshold effectively protects individuals with respiratory diseases. We speculate that NO2 was not considered robust in the sensitivity analysis due to its relatively low excess risk, which differs from findings in other studies [23]. This highlights the significant achievements of Shanghai’s air pollution control efforts. Similarly, CO, which did not pass the sensitivity analysis, exhibited a low excess risk in the exposure-response curve. Although its risk value was above zero before reaching the alert threshold, it showed only a brief increase before declining. Additionally, the maximum proportionate change in EACs associated with CO exposure did not exceed 3%. Consistent with our findings, other studies have also reported no significant association between CO concentrations and an increased risk of respiratory-related EACs [24].

In the stratified analysis, we found that individuals aged 65 and above experienced a higher exposure risk compared to other age groups, which is consistent with previous studies [25]. Additionally, we observed that the excess risk of EACs was higher during the daytime for all pollutants, which contrasts with findings from some studies [6]. This discrepancy may be attributed to Shanghai’s high nighttime humidity, which potentially alleviates pollution to some extent, as well as the tendency of people to stay indoors at night, reducing exposure. When plotting the exposure-response curves through stratified analysis, we observed that at single-day lags of 6 and 3 days, PM_2.5_ and O_3_ reached approximately 2.5% excess risk at their alert thresholds. Although this is not a particularly high value, it still suggests the need for a slight reduction in threshold levels to minimize respiratory disease-related EACs. Meanwhile, the alert threshold for PM_10_ appeared to be set too high, as its excess risk reached approximately 5% at the threshold on the 6th day of single-day lag in both spring and winter. Furthermore, during the 7-day cumulative lag in spring, the excess risk exceeded 7.5% before reaching the threshold. For SO2, the exposure-response curve exhibited an approximately linear increasing trend at a cumulative lag of 7 days in both spring and summer. This trend is supported by a study by Zhou et al. in Ganzhou, China, which demonstrated that during warm seasons, a 10 µg/m^3^ increase in SO2 concentration at a cumulative lag of 7 days was associated with a roughly 10% increase in emergency hospital admissions for respiratory diseases among individuals aged 65 and above [26]. Previous epidemiological studies have also linked SO2 exposure to adverse respiratory health effects, including impaired pulmonary function and increased mortality from respiratory diseases [27, 28]. The near-linear appearance of the exposure-response curve for SO2 in our study is likely due to the relatively narrow concentration range, giving the visual impression of linearity. In fact, since 2019, the daily average concentration of SO2 in Shanghai has not reached levels associated with hazardous effects on the exposure-response curve (Table 1). This highlights the outstanding achievements of Shanghai’s air pollution control efforts, demonstrating that SO2 mitigation has been a key priority for the government.

This study has several limitations. Firstly, we used daily average air pollution concentrations as a proxy for individual exposure levels. Although this approach may lead to some degree of exposure misclassification and potential underestimation of associations, it is a common limitation in most time-series studies [29, 30]. Secondly, due to the lack of final hospital admission clinical diagnosis data, we relied on pre-hospital diagnoses made by ambulance physicians for disease classification. However, given the professional expertise of on-site physicians and the relatively straightforward classification of respiratory diseases, this limitation is unlikely to introduce substantial bias in our results [31]. Thirdly, since we could not obtain daily pollutant data from individual monitoring stations or patients’ residential addresses, we relied on aggregated pollutant data from multiple national environmental monitoring stations rather than data from a single station. However, Shanghai is not an industrial city and lacks major pollution sources, and there were no significant differences in annual air pollutant concentrations across monitoring stations. Therefore, this approach is unlikely to introduce substantial measurement bias.

This study also has several strengths. Firstly, the Shanghai Emergency Medical Center (SEMC) provided high-quality data covering most hospitals in the city’s central districts. The large sample size and individual-level data enabled comprehensive statistical analysis, enhancing the validity of our findings. Secondly, with ongoing global climate change, defining seasons based solely on calendar months has become increasingly inaccurate in similar studies [32]. Instead, we applied a more standardized seasonal classification, allowing our study to more precisely capture the seasonal effects of different pollutants. Finally, we conducted a more refined stratified analysis to examine the associations between various pollutants and respiratory disease-related EACs. This approach allowed us to provide more precise threshold recommendations and offer evidence for the urgent need to control specific pollutants during different seasons.

Based on the *Air Quality Guidelines* (AQG) published by the World Health Organization and the actual pollution conditions in Shanghai, the local government should consider adjusting the recommended threshold for PM_10_ to align with the AQG levels during spring and winter. Additionally, the PM_2.5_ limit should be reduced by approximately one-third in spring and winter, while the ozone threshold should be lowered by about one-fourth in spring. Furthermore, considering the lag days associated with the highest risk values identified in this study, specific pollutant warnings should be issued to the public on consecutive pollution days to mitigate air pollution-related health risks. Our findings provide concrete policy recommendations that can assist emergency departments in optimizing ambulance deployment and help reduce the public’s potential exposure risks.

## 5. Conclusion

Our study identified the predominant lag periods and seasons during which different pollutants had the most pronounced impact on the risk of respiratory disease-related EACs. Sensitivity analysis confirmed the robustness of the model, revealing that this association was more significant among the elderly population and during daytime hours, with a notable seasonal effect. Local governments should consider adjusting the alert threshold for certain pollutants in specific seasons. Additionally, multi-regional collaborative studies could facilitate the reevaluation of national and international air pollution thresholds. Future research is expected to expand in both temporal and spatial scope, incorporating broader datasets to enhance understanding and policy development.

## Author Contributions

Conceptualization, H.J., Z.Z. (Zhifeng Zhang) and L.P.; methodology, H.J., Z.Z. (Zhifeng Zhang) and L.P.; software, W.L.; validation, Z.Z. (Zhifeng Zhang), L.P. and X.L.; formal analysis, H.J.; investigation, W.L.; resources, Z.Z. (Zhifeng Zhang); writing— original draft preparation, H.J.; writing—review and editing, H.J. and Z.Z. (Zhifeng Zhang); visualization, H.J.; supervision, Z.Z. (Zhifeng Zhang) and X.L.; project administration, Z.Z. (Zhifeng Zhang) and L.P.; funding acquisition, Z.Z. (Zhifeng Zhang).

## Funding

This research was funded by the Three-year action plan for strengthening the construction of the public health system in Shanghai, grant number GWVI-11.2-XD39.

## Institutional Review Board Statement

Not applicable

## Data Availability Statement

The datasets generated and/or analyzed during the current study are not publicly available owing to security protocols and privacy regulations, but they may be made available on reasonable request to the corresponding author.

## Conflicts of Interest

The authors declare no conflicts of interest.

## Notes

### Competing Interest Statement

The authors have declared no competing interest.

### Funding Statement

Yes

### Author Declarations

We received approval from the ethics committee of the Naval Medical University. All the participants voluntarily agreed to take part in our survey. During this study, the data provided by the Shanghai Emergency Medical Center had been thoroughly anonymized, with all directly identifiable personal information (such as names, ID numbers, and exact residential addresses) removed. Throughout the data collection and analysis phases, authors had no access to any information that could directly identify individual participants, nor could individual identities be inferred from the available data. Relevant files of ethics statement have been uploaded.

## References

1. Kumar, R.; Mrigpuri, P.; Sarin, R.; Saini, J. K.; Yadav, R.; Nagori, A.; Kabra, S. K.; Mukherjee, A.; Yadav, G., Air pollution and its effects on emergency room visits in tertiary respiratory care centers in Delhi, India. Monaldi Archives for Chest Disease 2024, 94 (1).

2. Li, Z. H.; Wang, X. M.; Liao, D. Q.; Zhang, Q.; Chen, Z. T.; Qiu, C. S.; Tang, X. L.; Li, H. M.; Du, L. Y.; Zhang, P. D.; Shen, D.; Zhang, X. R.; Gao, J.; Zhong, W. F.; Chen, P. L.; Huang, Q. M.; Song, W. Q.; Liu, D.; Li, C.; Chen, H.; Mao, C., Long-term air pollutants exposure and respiratory mortality: A large prospective cohort study. Ecotoxicol Environ Saf 2024, 274, 116176.

3. Ren, M.; Li, N.; Wang, Z.; Liu, Y.; Chen, X.; Chu, Y.; Li, X.; Zhu, Z.; Tian, L.; Xiang, H., The short-term effects of air pollutants on respiratory disease mortality in Wuhan, China: comparison of time-series and case-crossover analyses. Scientific Reports 2017, 7 (1), 40482.

4. Monoson, A.; Schott, E.; Ard, K.; Kilburg-Basnyat, B.; Tighe, R. M.; Pannu, S.; Gowdy, K. M., Air pollution and respiratory infections: the past, present, and future. Toxicol Sci 2023, 192 (1), 3–14.

5. Wang, Y.-C.; Lin, Y.-K.; Chen, Y.-J.; Hung, S.-C.; Zafirah, Y.; Sung, F.-C., Ambulance Services Associated with Extreme Temperatures and Fine Particles in a Subtropical Island. Scientific Reports 2020, 10 (1), 2855.

6. Chen, T.-T.; Zhan, Z.-Y.; Yu, Y.-M.; Xu, L.-J.; Guan, Y.; Ou, C.-Q., Effects of hourly levels of ambient air pollution on ambulance emergency call-outs in Shenzhen, China. Environmental Science and Pollution Research 2020, 27, 24880–24888.

7. Cui, X.; Tian, Y.; Yin, Z.; Huang, S.; Yin, P., The Impact of Ambient PM2.5 on Emergency Ambulance Dispatches Due to Circulatory System Disease Modified by Season and Temperature in Shenzhen, China. Atmosphere 2025, 16 (2), 198.

8. Zhou, Q.; Shi, H.; Wu, R.; Zhu, H.; Qin, C.; Liang, Z.; Sun, S.; Zhao, J.; Wang, Y.; Huang, J., Associations between hourly ambient particulate matter air pollution and ambulance emergency calls among 3,022,164 patients: time stratified case-crossover study. JMIR Public Health and Surveillance 2023.

9. World Health, O., WHO global air quality guidelines: particulate matter (PM2.5 and PM10), ozone, nitrogen dioxide, sulfur dioxide and carbon monoxide. World Health Organization: Geneva, 2021.

10. Sun, Y.; Milando, C. W.; Spangler, K. R.; Wei, Y.; Schwartz, J.; Dominici, F.; Nori-Sarma, A.; Sun, S.; Wellenius, G. A., Short term exposure to low level ambient fine particulate matter and natural cause, cardiovascular, and respiratory morbidity among US adults with health insurance: case time series study. bmj 2024, 384.

11. China, M. o. E. a. E. o. t. P. s. R. o. Ambient air quality standards. Available on https://www.mee.gov.cn/ywgz/fgbz/bz/bzwb/dqhjbh/dqhjzlbz/201203/t20120302_224165.shtml.

12. Administration, C. M. Division of climatic seasons. Available on https://www.cma.gov.cn/zfxxgk/gknr/flfgbz/bz/202307/t20230712_5642647.html.

13. Wu, W.; Chen, B.; Wu, G.; Wan, Y.; Zhou, Q.; Zhang, H.; Zhang, J., Increased susceptibility to temperature variation for non-accidental emergency ambulance dispatches in Shenzhen, China. Environmental Science and Pollution Research 2021, 28 (24), 32046–32056.

14. Zhou, Y.; Jin, Y.; Zhang, Z., Short-term exposure to various ambient air pollutants and emergency department visits for cause-stable ischemic heart disease: a time-series study in Shanghai, China. Scientific Reports 2023, 13 (1), 16989.

15. Ho, A. F. W.; Hu, Z.; Woo, T. Z. C.; Tan, K. B. K.; Lim, J. H.; Woo, M.; Liu, N.; Morgan, G. G.; Ong, M. E. H.; Aik, J., Ambient air quality and emergency hospital admissions in Singapore: a time-series analysis. International Journal of Environmental Research and Public Health 2022, 19 (20), 13336.

16. Kim, H.; Yu, W., Single-day and cumulative effects of ambient particulate matter exposure on emergency department visits for respiratory disease in South Korea. Hong Kong Journal of Emergency Medicine 2022, 29 (1), 17–23.

17. Liu, Y.; Wang, Y.; Dong, J.; Wang, J.; Bao, H.; Zhai, G., Association between air pollution and emergency department visits for upper respiratory tract infection inLanzhou, China. Environmental Science and Pollution Research 2022, 29 (19), 28816–28828.

18. Shi, H.; Zhou, Q.; Zhang, H.; Sun, S.; Zhao, J.; Wang, Y.; Huang, J.; Jin, Y.; Zheng, Z.; Wu, R.; Zhang, Z., The Combined Effects of Hourly Multi-Pollutant on the Risk of Ambulance Emergency Calls: A Seven-Year Time Series Study. Toxics 2023, 11 (11), 895.

19. Huang, H.; Lin, C.; Liu, X.; Zhu, L.; Avellán-Llaguno, R. D.; Lazo, M. M. L.; Ai, X.; Huang, Q., The impact of air pollution on COVID-19 pandemic varied within different cities in South America using different models. Environmental Science and Pollution Research 2022, 29 (1), 543–552.

20. Tang, C.; Chen, Y.; Song, Q.; Ma, J.; Zhou, Y.; Gong, L.; Chen, X.; Qu, J.; Luo, Y., Short-term exposure to air pollution and occurrence of emergency stroke in Chongqing, China. International Archives of Occupational and Environmental Health 2021, 94, 69–76.

21. Ma, Y.; Shen, J.; Zhang, Y.; Wang, H.; Li, H.; Cheng, Y.; Guo, Y., Short-term effect of ambient ozone pollution on respiratory diseases in western China. Environmental Geochemistry and Health 2022, 44 (11), 4129–4140.

22. Peng, J.; Chen, J.; Wu, X.; Qian, J.; Li, N.; Yi, Y.; Huang, Y.; Lu, J.; Zhang, W.; Li, Z.; Li, Z.; Li, M.; Liu, X., Short-term effects of low-level PM2.5, PM10, O3, and tropical meteorological conditions on emergency department visits for respiratory diseases in Haikou, China. Asian Pacific Journal of Tropical Medicine 2024, 17 (7), 317–328.

23. Fu, Y.; Zhang, W.; Li, Y.; Li, H.; Deng, F.; Ma, Q., Association and interaction of O3 and NO2 with emergency room visits for respiratory diseases in Beijing, China: a time-series study. BMC Public Health 2022, 22 (1), 2265.

24. Wang, X.; Leng, M.; Liu, Y.; Qian, Z.; Zhang, J.; Li, Z.; Sun, L.; Qin, L.; Wang, C.; Howard, S. W.; Vaughn, M. G.; Yan, Y.; Lin, H., Different sized particles associated with all-cause and cause-specific emergency ambulance calls: A multicity time-series analysis in China. Science of The Total Environment 2021, 783, 147060.

25. Johnston, F. H.; Salimi, F.; Williamson, G. J.; Henderson, S. B.; Yao, J.; Dennekamp, M.; Smith, K.; Abramson, M. J.; Morgan, G. G., Ambient Particulate Matter and Paramedic Assessments of Acute Diabetic, Cardiovascular, and Respiratory Conditions. Epidemiology 2019, 30 (1), 11–19.

26. Zhou, X.; Gao, Y.; Wang, D.; Chen, W.; Zhang, X., Association Between Sulfur Dioxide and Daily Inpatient Visits With Respiratory Diseases in Ganzhou, China: A Time Series Study Based on Hospital Data. Frontiers in Public Health 2022, 10.

27. Chen, J.; Shi, C.; Li, Y.; Ni, H.; Zeng, J.; Lu, R.; Zhang, L., Effects of short-term exposure to ambient airborne pollutants on COPD-related mortality among the elderly residents of Chengdu city in Southwest China. Environmental health and preventive medicine 2021, 26, 1–10.

28. Orellano, P.; Reynoso, J.; Quaranta, N., Short-term exposure to sulphur dioxide (SO2) and all-cause and respiratory mortality: A systematic review and meta-analysis. Environment international 2021, 150, 106434.

29. Shin, H. H.; Maquiling, A.; Thomson, E. M.; Park, I.-W.; Stieb, D. M.; Dehghani, P., Sex-difference in air pollution-related acute circulatory and respiratory mortality and hospitalization. Science of the Total Environment 2022, 806, 150515.

30. Dionisio, K. L.; Chang, H. H.; Baxter, L. K., A simulation study to quantify the impacts of exposure measurement error on air pollution health risk estimates in copollutant time-series models. Environmental Health 2016, 15, 1–10.

31. Mould-Millman, C. N.-K., Assessing use of the South African triage scale in the Western Cape government emergency medical services system. 2022.

32. Zhou, Q.; Shi, H.; Wu, R.; Zhu, H.; Qin, C.; Liang, Z.; Sun, S.; Zhao, J.; Wang, Y.; Huang, J.; Jin, Y.; Zheng, Z.; Li, J.; Zhang, Z., Associations Between Hourly Ambient Particulate Matter Air Pollution and Ambulance Emergency Calls: Time-Stratified Case-Crossover Study. JMIR Public Health Surveill 2023, 9, e47022.

